# Feasibility of Noninvasive Atrial and Ventricular Activation, Conduction-Velocity, and Site-of-Origin Mapping with Solid-State Magnetocardiography: A Preclinical Validation Study

**DOI:** 10.64898/2026.07.08.26357590

**Authors:** Kelly A. Brennan, Sabyasachi Bandyopadhyay, Charles Sillett, Jennifer K. Lyons, Makoto Kameno, Yasushi Terazono, Prasanth Ganesan, Xichong Liu, Gentaro Ikeda, Hiroyuki Takashima, Yuka Matsuura, Mariko Koike-Ieki, Phillip C. Yang, Miguel Rodrigo, Paul J. Wang, Sanjiv M. Narayan, Albert J. Rogers

## Abstract

**Background:** Characterizing cardiac activation by its site of origin, propagation, and conduction velocity underlies arrhythmia diagnosis and management, but invasive electrophysiology (EP) mapping requires vascular access, fluoroscopy, and sedation. Magnetocardiography (MCG) enables contactless mapping, and recent solid-state sensors remove the cost, cryogenic, and shielding barriers of legacy systems. We assessed the feasibility of a novel solid-state MCG system for noninvasive arrhythmia site-of-origin (SOO) localization and activation reconstruction, benchmarked against electrocardiographic imaging (ECGi).

**Methods:** In nine swine implanted with right atrial and right ventricular pacing leads, we recorded MCG and ECGi simultaneously during atrial and ventricular pacing. Invasive epicardial contact EP mapping provided the activation-time reference and MRI-derived lead-tip location the SOO reference. Local activation time (LAT), conduction velocity (CV), and SOO were compared on a co-registered chamber mesh. SOO error was the Euclidean distance to the MRI lead tip; LAT and CV agreement with EP were quantified by Pearson r and compared using Wilcoxon signed-rank tests.

**Results:** Across 17 datasets (8 atrial, 9 ventricular), median SOO error was lower for MCG than ECGi in the atrium (19.6 vs 31.2 mm; p=0.023) and ventricle (12.0 vs 26.1 mm; p=0.074). LAT agreement with EP was comparable between modalities and higher in the ventricle (MCG r=0.63; ECGi r=0.68) compared with the atrium (MCG r=0.40; ECGi r=0.53), each correlating with invasive EP mapping above chance. CV agreement was modest and numerically higher for MCG in the ventricle.

**Conclusions:** Solid-state MCG was feasible for noninvasive site-of-origin localization and activation mapping, with accuracy comparable to ECGi, motivating larger prospective studies to define its clinical role in noninvasive mapping.

**Clinical Perspective:** *What Is Known:*

- Invasive electrophysiology mapping is the reference standard for localizing the arrhythmia site of origin and characterizing cardiac activation, but it requires vascular access, fluoroscopy, and prolonged sedation within the electrophysiology laboratory.
- Existing noninvasive options have important limitations: electrocardiographic imaging depends on concomitant thoracic imaging, size-dependent vest fit, and torso-conductivity modeling, while legacy magnetocardiography has required costly cryogenic and magnetically shielded infrastructure.

*What the Study Adds:*

- In a preclinical large-mammal model referenced to invasive electrophysiology and MRI, a novel contactless solid-state magnetocardiography system feasibly localized atrial and ventricular sites of origin and reconstructed activation without body-surface electrodes.
- Magnetocardiography localized the site of origin more accurately than ECGi in the atrium, and showed numerically lower localization error in the ventricle.
- Magnetocardiography displayed activation-time and conduction-velocity agreement with invasive electrophysiology comparable to electrocardiographic imaging in both chambers.
- Solid-state magnetocardiography may offer a scalable, noninvasive approach for preprocedural localization and complementary activation mapping, motivating larger prospective clinical validation.

## Introduction

Characterizing cardiac activation, specifically where it originates, how it propagates, and how fast it conducts,^1^ underlies both the physiological study and clinical management of atrial and ventricular arrhythmias, directly informing catheter ablation planning, procedure risk assessment, mechanistic phenotyping, and emerging noninvasive therapies.^2,3^ Invasive electrophysiology mapping is the reference standard, but requires vascular access, fluoroscopic guidance, prolonged sedation, and time within the electrophysiology laboratory.^2^ These costs are amplified in patients in whom invasive mapping is technically constrained, hemodynamically risky, or poorly tolerated.^2,4^ A practical, accurate, noninvasive method to reconstruct activation and localize the arrhythmia site-of-origin (SOO) before the procedure would therefore have value across both catheter-based and noninvasive arrhythmia therapy.

Of the available noninvasive mapping technologies, electrocardiographic imaging (ECGi) has the longest track record in clinical practice. ECGi combines body-surface potentials from a dense electrode arrangement with a patient-specific heart-torso geometry derived from thoracic computed tomography or magnetic resonance imaging (MRI) to reconstruct epicardial activation, and has been applied in adult and selected pediatric populations, including patients with Wolff-Parkinson-White syndrome and congenital heart disease.^5–7^ However, ECGi is constrained by the need for concomitant cross-sectional thoracic imaging, size-dependent vest fitting and skin adhesive, and sensitivity of signal reconstruction to inter-organ conductivity.^5,6^

In contrast, magnetocardiography (MCG) captures both tangential and volumetric current activity without body-surface electrodes. This contactless approach reduces reliance on high-fidelity torso modeling and provides a complementary view to ECGi,^8,9^ potentially improving spatial characterization and enabling longer recording durations. Traditional MCG systems based on superconducting quantum interference devices or optically pumped magnetometers have remained outside routine pediatric and adult arrhythmia practice because of cost, magnetic-shielding requirements, and cryogenic infrastructure.^9,10^ Recent advances in solid-state magnetic sensor technology have produced compact MCG systems that record without these limitations, lowering the infrastructure barrier to clinical deployment and opening the possibility of a contactless, scalable noninvasive mapping modality suitable for adult, pediatric, and longitudinal use.^10–12^

The objective of this study was to assess the feasibility of a novel solid-state MCG system for noninvasive characterization of cardiac activation, specifically local activation time (LAT), conduction velocity (CV), and site of origin (SOO), and to benchmark its performance against ECGi. Invasive epicardial EP mapping and MRI-derived pacing-lead location served as the reference standards. We hypothesized that (1) MCG-derived activation-time and conduction velocity maps would correlate with the invasive EP reference above chance, and (2) that this agreement, and site-of-origin localization accuracy, would be comparable to ECGi.

## Methods

### Study Design and Cohort

We evaluated the feasibility of a novel solid-state MCG system (TDK Corporation, Tokyo, Japan) for arrhythmia SOO localization and activation mapping. Pacing lead implantation was performed in a cohort of nine swine (mean weight: 43.4 ± 6.3 kg, range 34–51 kg; 6 female, 3 male), enabling controlled chamber-level pacing. SOO was determined using the MRI-derived lead localization by a board-certified electrophysiologist. To contextualize MCG performance, comparative analyses were conducted with ECGi. See **Figure 1** for an overview of the study procedure.

**Figure 1.**
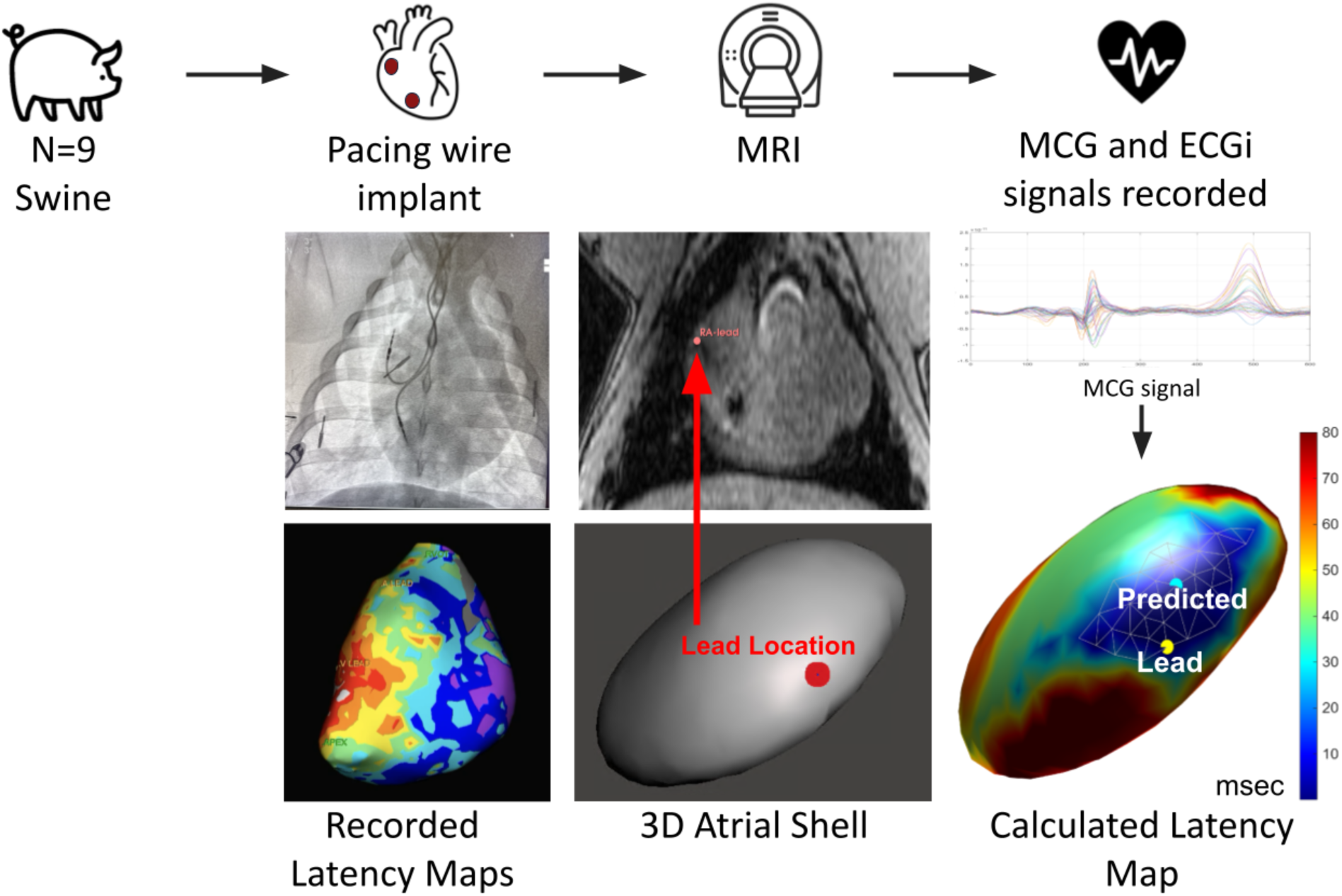
End-to-end study workflow for comparative evaluation of MCG and ECGi against invasive electrophysiology (EP) and MRI references. In nine swine, right atrial and right ventricular pacing leads were implanted, after which MRI was acquired for chamber segmentation and lead-tip localization. Simultaneous MCG and ECGi signals were then recorded during chamber-level pacing, and invasive epicardial EP contact mapping provided the activation-time reference standard. For each modality, local activation time (LAT) maps were reconstructed and co-registered to a common chamber mesh in a shared coordinate frame; the estimated site-of-origin (SOO; earliest-activation centroid) was compared with the MRI-derived pacing lead tip, and MCG- and ECGi-derived activation-time and conduction-velocity maps were compared with the EP reference. Bottom panels (left to right) show a representative recorded EP activation map, MRI-based lead-tip identification with the corresponding 3D chamber shell, and a calculated activation map with the predicted SOO and the reference lead. Atrial and ventricular analyses followed the same pipeline. Localization error (Euclidean distance from the modality-specific SOO to the MRI lead-tip reference) and activation-time and conduction-velocity agreement with invasive EP were the primary endpoints. MCG, magnetocardiography; ECGi, electrocardiographic imaging; MRI, magnetic resonance imaging; EP, electrophysiology; SOO, site of origin; LAT, local activation time.

A total of eighteen chamber-level activation maps were collected, equally distributed between atrial and ventricular datasets. One atrial dataset was excluded for absence of atrial capture, leaving 8 atrial and 9 ventricular datasets analyzed.

Animal care and ethics approvals were obtained under Stanford University protocol 34498. The data that support the findings of this study are available from the corresponding author upon reasonable request.

### Pacing Protocol and Reference Standards

Pacing lead implantation was performed under sterile conditions using percutaneous right and left internal jugular vein access established via the Seldinger technique. After local anesthesia and ultrasound-guided venipuncture, a guidewire was advanced into the right atrium and exchanged for 7F introducer sheaths. Through the sheaths, two MRI-conditional pacing wires (model 4076, Medtronic, Minneapolis, MN) were sequentially positioned under fluoroscopic guidance. One pacing lead was placed in the right atrium and one in the right ventricle, targeting the free wall or appendage of the right atrium and the free wall or right ventricular outflow tract of the right ventricle. Lead placement was confirmed by fluoroscopy and pacing-threshold testing to ensure stable capture in both chambers.

Invasive epicardial contact electrophysiology mapping served as the reference standard for activation-sequence characterization and SOO determination, enabling quantitative comparison with both MCG and ECGi reconstructions; the mapping procedure is detailed below (see Epicardial Reference Mapping and Harmonization).

### Imaging, Segmentation, and Mesh Generation

Following pacing lead implantation, MRI was performed to establish a reference standard for anatomical localization. This volumetric imaging was performed by 3T MRI (Signa 3T-HDx, GE, USA) using TIR Body coil and cardiac vector gating. Following localizer images, a high-resolution 3D gradient-echo sequence (3D fast gradient echo; ECG-gated, echo time minimum, flip angle 15°, slice thickness 2 mm, matrix 320×256, field of view 40 cm) was acquired. The imaging protocol was designed to balance spatial resolution with coverage, ensuring complete visualization of fiducials, cardiac anatomy, and lead artifacts.

Identical MR-visible skin fiducials were used for both MCG and ECGi modalities and distinguished by anatomical placement. For ECGi, 50 fiducials were arranged as 10 vertical strips of 5 markers each, distributed circumferentially around the torso to co-register the body-surface electrode array. For MCG, a set of 5 fiducials was placed over the cardiac silhouette to register the heart’s location to the MCG sensor system. All fiducials were included in the imaging volume to enable accurate cross-modality registration.

Lead localization was performed by a board-certified electrophysiologist who identified susceptibility artifacts from the pacing wires, which allowed clear delineation of lead-tip positions. These data, combined with cardiac chamber geometry and torso surface contours, defined a unified reference framework for subsequent comparisons with noninvasive mapping results.

Cardiac chamber segmentation and surface mesh generation were performed using semi-automated methods with manual refinement. Segmentation was conducted in 3D Slicer (version 5.10.0), with volumes reviewed against source images to confirm anatomical accuracy. Atrial and ventricular geometries were separated along the atrioventricular plane. Surface post-processing, including smoothing and mesh repair, was performed in Autodesk Meshmixer (version 3.5), generating closed, watertight geometries suitable for inverse mapping and cross-modality registration.

### MCG and ECGi Data Acquisition and Reconstruction

Simultaneous MCG and ECGi recordings were performed during controlled atrial and ventricular pacing to ensure consistent activation origin across modalities. Pacing was delivered via implanted leads using chamber-specific overdrive protocols. For atrial pacing, cycle lengths were set at 10–20% above baseline sinus rhythm rate to ensure stable capture, minimize fusion, and maintain hemodynamic stability. Ventricular pacing was performed at programmed cycle lengths selected using similar principles. Pacing sequences were delivered and acquired continuously over 10-minute intervals for each condition. Pacing output was minimized (typically ∼1 mA) to reduce far-field capture and pacing artifact.

ECGi data were acquired using the EnSite Precision NavX system (Abbott Laboratories, Minneapolis, MN) via a custom non-ferromagnetic recording configuration to enable simultaneous acquisition with MCG. A total of 50 nonferrous adhesive electrodes (Vitrode V, Ag/AgCl adhesive gel electrodes with lead wires; Nihon Kohden, Tokyo, Japan) were circumferentially distributed around the thorax to provide body-surface potential coverage. Signals were recorded continuously and processed to generate ECGi-derived activation maps using standard inverse reconstruction techniques.^13^

MCG recordings were obtained concurrently using a solid-state magnetic sensor array positioned external to the thorax without direct subject contact. Data were collected over extended recording windows (at least 10 minutes), enabling signal averaging and improved signal-to-noise characteristics. MCG signals were processed to derive magnetic field maps and reconstructed into cardiac activation patterns using inverse modeling approaches aligned to subject-specific anatomical geometry.

Quality control procedures were applied to ensure analyzability of recordings. These included verification of consistent pacing capture, absence of fusion beats, signal stability over the acquisition window, and adequate signal-to-noise ratio for both MCG and ECGi datasets. Recordings not meeting these criteria were either repeated or excluded from downstream analysis.

### Epicardial Reference Mapping and Co-registration

Epicardial contact electrophysiology mapping was performed across the entire heart under atrial, ventricular, and sinus rhythm pacing modalities. Mapping utilized the HD Grid catheter in conjunction with the EnSite Precision NavX system (Abbott Laboratories, Minneapolis, MN), delivered via a subxyphoid approach to access the epicardial space.

Local activation time maps were extracted from EP studies for atrial and ventricular pacing datasets after beat-averaging under stable rhythm conditions. To place EP data into a common anatomical frame, EP sinus-rhythm geometry was co-registered to the sinus atrial and ventricular shells derived from MRI segmentation. Registration used chamber-specific lead fiducials so that right atrial and right ventricular lead reference points from MRI were aligned to the corresponding right atrial and right ventricular fiducials from the EP study, defined from impedance-derived pacing wire locations.

After co-registration, the MCG and ECGi activation maps were brought into vertex-wise correspondence with the EP reference mesh in the shared coordinate frame by rigid (iterative closest point) alignment followed by nearest-point matching. Chamber-specific flat-region trimming and finite-value masking, computed on the EP reference mesh, then restricted the analysis to reliable surface vertices, enabling consistent vertex-wise comparison across EP, MCG, and ECGi. The resulting aligned EP LAT maps were then used for downstream analyses of activation pattern characterization, conduction velocity, and site-of-origin localization.

### Site-of-Origin Localization

Site-of-origin (SOO) localization was performed separately for atrial and ventricular pacing datasets after modality harmonization onto a common chamber mesh representation. For each modality (MCG, ECGi, and epicardial contact LAT map), local activation time values were first restricted to analyzable vertices using the chamber-specific flat-region trim mask (**Supplement 2**), and only finite LAT values were retained. Candidate earliest-activation vertices were then identified by selecting the lowest-LAT subset under a prespecified rule (fixed early-point count or equivalent percentile threshold, prespecified before final analysis lock; **Supplement 3**).

To improve robustness against isolated noise points, the candidate earliest vertices were constrained using mesh connectivity. A vertex adjacency graph was derived from triangular faces, and connected components were computed on the earliest-activation subset. The primary SOO cluster was defined as the dominant component according to prespecified criteria (largest component with lowest central LAT tendency). Clusters of fewer than two mesh-connected vertices were treated as spurious, and the largest connected component among the earliest-activation candidates was retained as the site of origin.

The SOO coordinate for each modality was defined as the 3-dimensional centroid of the retained SOO cluster vertices in the shared mesh coordinate frame. This definition yields a stable point estimate that is less sensitive to single-vertex outliers than minimum-value index selection alone. When needed, supporting descriptors (cluster size, spatial spread, and centroid dispersion) were recorded for quality control and sensitivity analyses.

Localization accuracy was quantified by Euclidean distance from each modality-specific SOO coordinate to the MRI-derived pacing lead reference coordinate in the same chamber and coordinate frame. Distances were reported in millimeters for atrial and ventricular analyses separately. For modality comparison, paired per-dataset differences (MCG minus ECGi distance-to-lead) were computed so that negative values indicated lower localization error for MCG.

### Activation-Time and Conduction-Velocity Agreement

Prior to agreement analysis, LAT fields for all modalities (MCG, ECGi, and the EP reference) were spatially smoothed with a chamber-specific Laplacian kernel matched to the chambers’ spatial resolution to model a continuous activation wavefront. Rather than choosing the smoothing strength ad hoc, the number of Laplacian iterations was calibrated per chamber: using an impulse-response characterization of the relationship between iteration count and the kernel’s Gaussian-equivalent spatial scale (σ) at a fixed step size (λ = 0.5), the iteration count was set so that σ matched each chamber’s spatial resolution. For the ventricle, σ was set to the median MCG site-of-origin localization error (approximately 12 mm). For the atrium, whose median localization error was larger (approximately 20 mm), the kernel at this value over-smoothed the comparatively small atrial maps, so σ was instead capped at approximately 12 mm, matching the ventricular scale. Because the iteration-to-σ mapping depends on mesh vertex spacing, these targets corresponded to 9 smoothing iterations for atrial maps and 7 for ventricular maps, with the identical chamber-specific kernel applied to MCG, ECGi, and the EP reference.

Low, gap-disconnected baseline outliers in the EP maps (stray near-zero vertices separated by more than 10ms from the activation distribution) were excluded so that they did not anchor the zero-reference. Activation-time agreement with EP was quantified by Pearson (r) and Spearman (ρ) correlation, and vertex-wise conduction velocity was estimated as the magnitude of a local least-squares plane-fit gradient of the smoothed LAT field and compared with EP by Pearson r.

### Statistical Analysis

The analysis was prespecified around three endpoint families: (1) activation-time (LAT) agreement, (2) conduction-velocity agreement, and (3) SOO localization accuracy. Activation-time and conduction-velocity agreement were evaluated for MCG vs EP and ECGi vs EP within each chamber using Pearson correlation coefficient (*r*); the Spearman rank-correlation coefficient (ρ) was additionally reported, and vertex-wise conduction velocity was compared with EP analogously using the plane-fit gradient of the smoothed LAT field. SOO localization endpoints were defined as distance-to-lead error (mm) relative to MRI lead reference coordinates for atrial and ventricular pacing datasets.

All primary analyses were chamber-stratified (atrial, ventricular), with modality contrasts performed within paired datasets. Localization error (distance-to-lead) was summarized as median [IQR], whereas activation-time and conduction-velocity agreement (Pearson r and Spearman ρ) were summarized as mean ± standard deviation with nonparametric bootstrap 95% confidence intervals. For SOO localization error, paired MCG and ECGi distance-to-lead values were compared within each chamber using the Wilcoxon signed-rank test. For activation-time and conduction-velocity agreement, we additionally tested whether each modality’s correlation with EP exceeded zero (one-sample Wilcoxon signed-rank test). Paired differences are reported with bootstrap 95% confidence intervals (10,000 resamples) and the matched-pairs rank-biserial correlation as the effect size. A two-sided α of 0.05 was used without multiplicity adjustment given the feasibility scope. Because the Wilcoxon test is conservative at this sample size, interpretation emphasizes effect estimates and confidence intervals over p-values.

## Results

### SOO Localization Accuracy Versus MRI Lead Location

Across the 17 analyzable chamber-level datasets (8 atrial, 9 ventricular; one atrial dataset excluded as described in the Study Design and Cohort), SOO was calculated by each method in each chamber. Representative activation maps with earliest activation sites are shown in **Figure 2**. Localization error distributions differed by chamber and modality (**Table 1**, **Figure 3**). In atrial pacing datasets, median distance-to-lead error was 19.6 mm [15.6, 22.9] for MCG and 31.2 mm [20.1, 46.0] for ECGi. The paired atrial difference (MCG minus ECGi) had a median of -12.1 mm, indicating significantly lower localization error for MCG (Wilcoxon signed-rank p=0.023; n=8).

**Figure 2.**
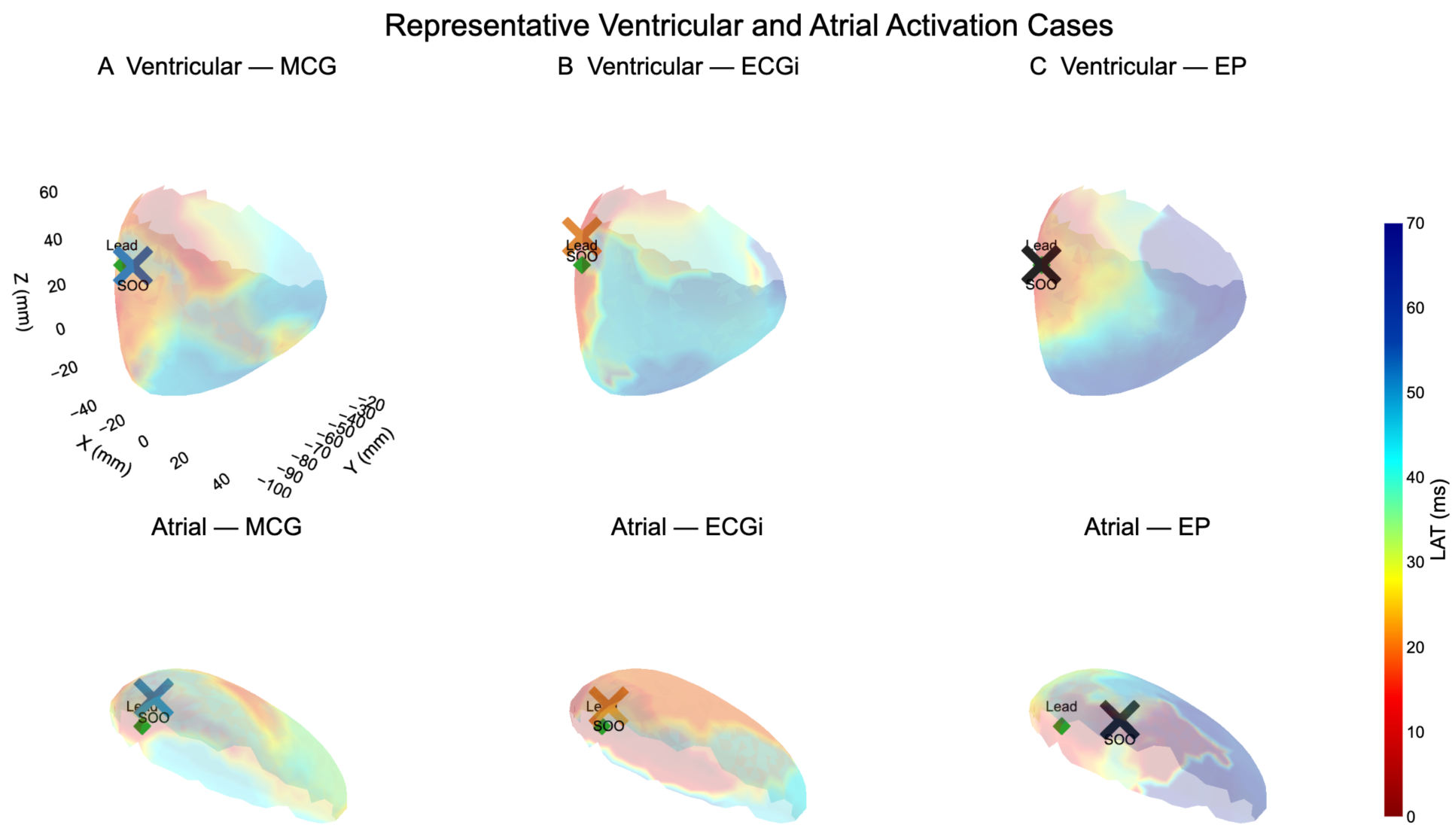
Representative ventricular (top row) and atrial (bottom row) cases comparing noninvasive MCG and ECGi-derived site-of-origin (SOO) estimates against the invasive EP and MRI reference in a shared chamber coordinate frame. Columns show MCG, ECGi, and the epicardial EP reference, with all panels rendered from the same viewpoint. The translucent chamber surface is colored by local activation time (LAT; scale at right), with earliest activation in red. In each panel, the green diamond marks the MRI-derived pacing lead-tip reference (Lead) and the X marks the modality-specific estimated SOO centroid (the centroid of the earliest-activation cluster): blue for MCG, orange for ECGi, and black for EP. Closer proximity of the SOO marker to the Lead indicates more accurate localization. This figure illustrates case-level concordance and discordance patterns interpreted qualitatively alongside the quantitative distance-to-lead metrics (Table 1, Figure 3).

**Figure 3.**
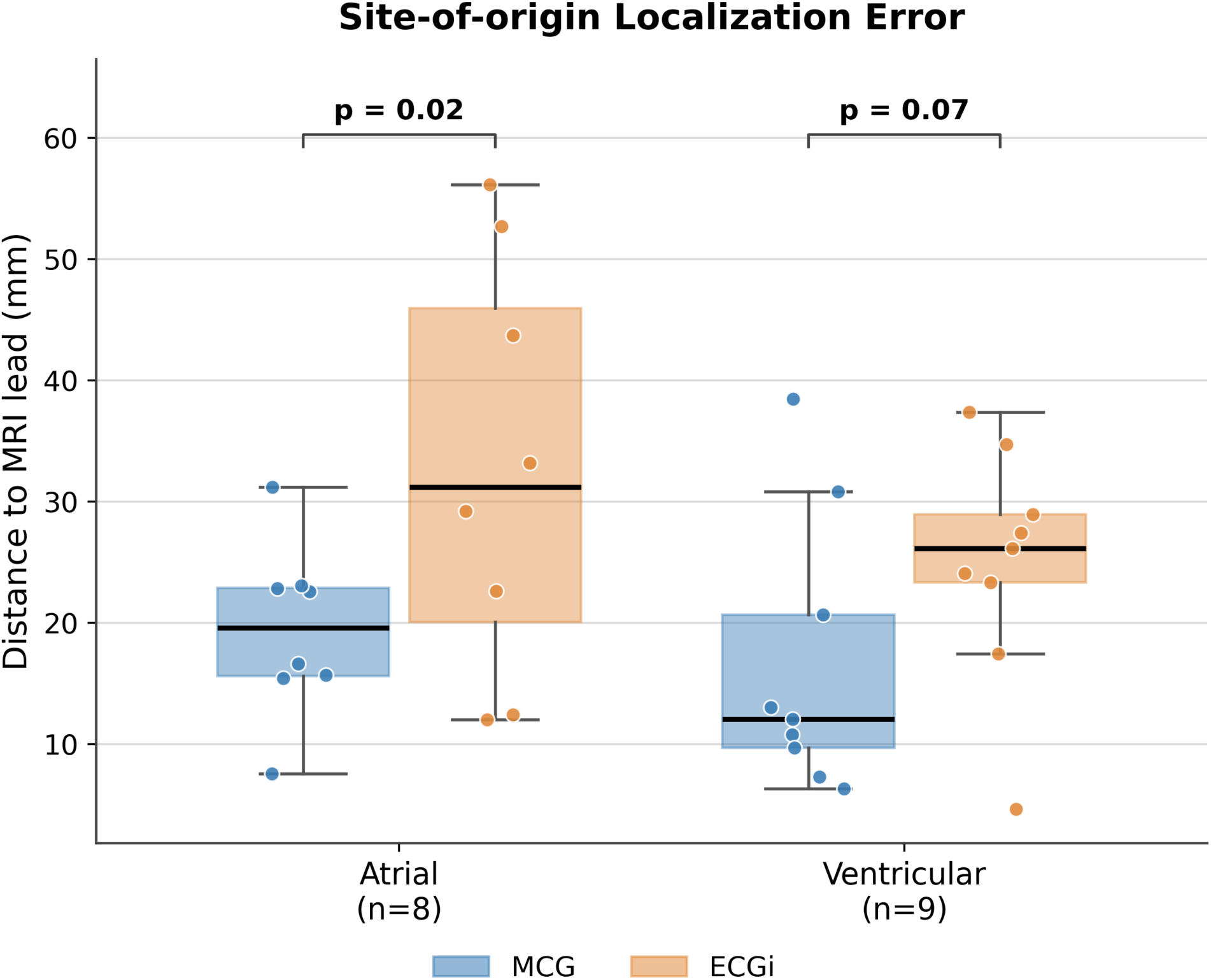
Site-of-origin localization error by chamber and modality, quantified as the Euclidean distance from each modality’s estimated SOO to the MRI-derived pacing lead tip (mm). Boxes show the median (heavy line) and interquartile range, whiskers extend to 1.5× the interquartile range, and individual study values are overlaid as points (atrial n=8, ventricular n=9; MCG in blue, ECGi in orange). Brackets indicate the paired MCG-versus-ECGi contrast (two-sided Wilcoxon signed-rank): p=0.02 in the atrium and p=0.07 in the ventricle. Lower values indicate localization closer to the reference lead; MCG error was lower than ECGi in both chambers, reaching statistical significance in the atrium. Values correspond to Table 1.

**Table 1:**
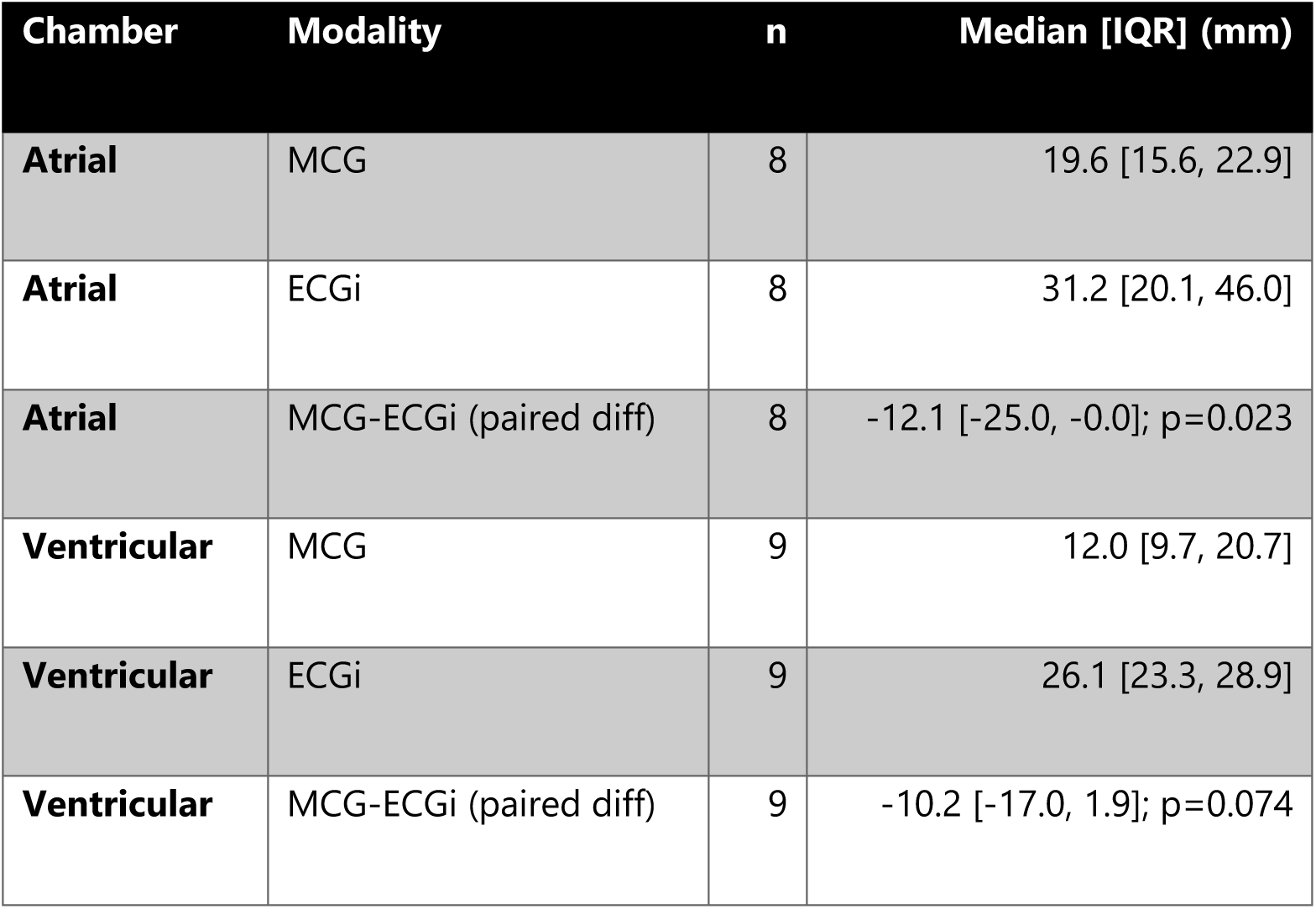
Distance metric is the Euclidean distance from modality-specific SOO centroid to MRI-derived lead-tip reference in the co-registered chamber mesh coordinate frame. Values for each modality are the median [interquartile range] of the per-study distance-to-lead. The paired MCG-minus-ECGi difference is reported as the median difference with its nonparametric bootstrap 95% confidence interval (10,000 resamples) and was tested by the Wilcoxon signed-rank test (p shown in the paired-difference row). Atrial analyses include 8 datasets; ventricular include 9.

In ventricular pacing datasets, median distance-to-lead error was 12.0 mm [9.7, 20.7] for MCG and 26.1 mm [23.3, 28.9] for ECGi. The paired ventricular difference (MCG minus ECGi) had a median of -10.2 mm, favoring lower localization error for MCG (Wilcoxon signed-rank p=0.074; n=9). Representative case-level localization overlays are shown in Figure 2.

### LAT Agreement and Conduction Velocity

LAT agreement with EP reference maps is summarized in **Table 2** and **Figure 4**. Atrial agreement (n=8) was lower and comparable between modalities: MCG vs EP mean Pearson r 0.40 ± 0.39 (Spearman ρ 0.40) and ECGi vs EP mean Pearson r 0.53 ± 0.23 (ρ 0.53); the paired difference was not significant (Wilcoxon signed-rank p=0.38). Each modality correlated with EP above chance (MCG p=0.039; ECGi p=0.008).

**Figure 4.**
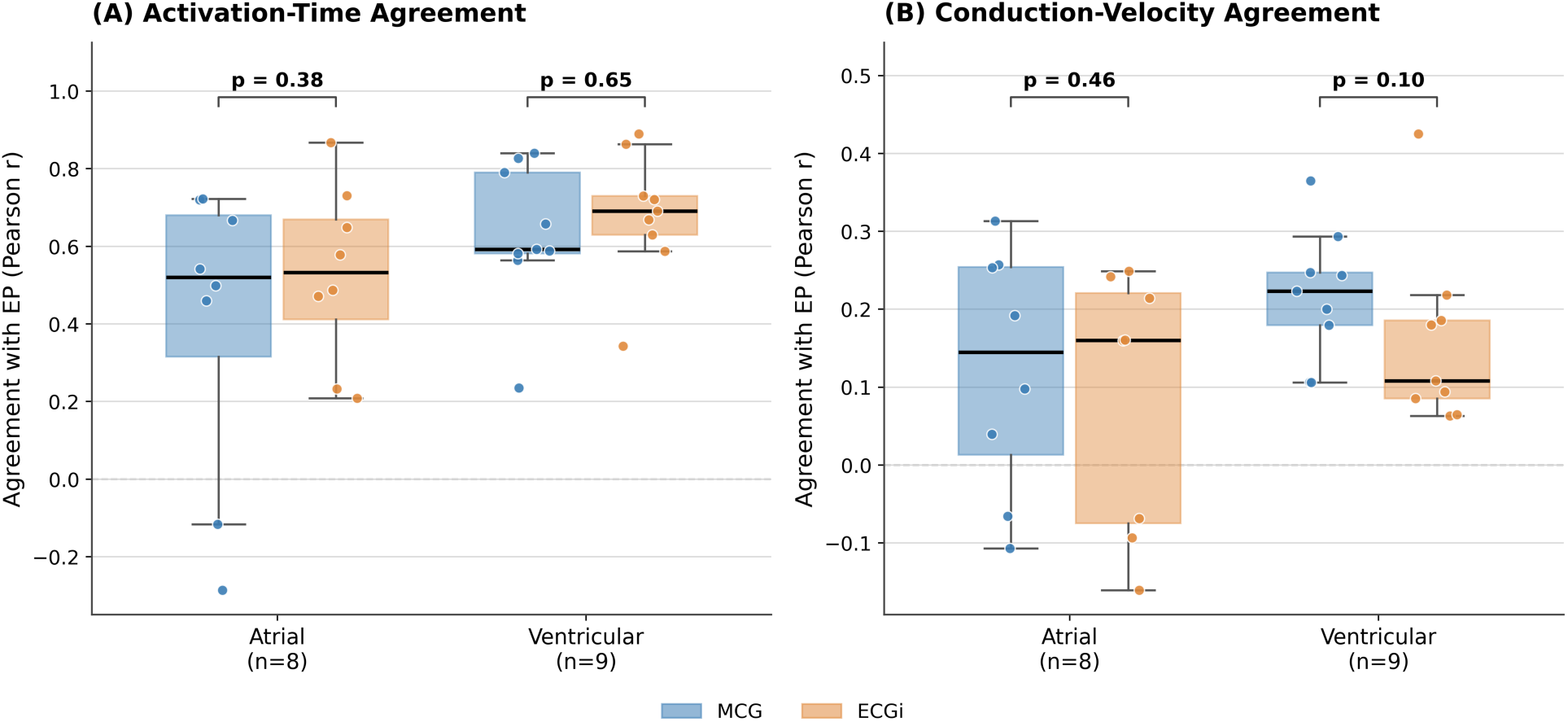
Chamber-stratified agreement between noninvasive reconstruction (MCG, blue; ECGi, orange) and the invasive EP reference, quantified as the per-study Pearson correlation (r) with EP. **(A)** Activation-time agreement, computed on spatially smoothed, zero-referenced LAT maps over the trimmed analyzable surface. **(B)** Conduction-velocity agreement, computed as the Pearson correlation between each modality’s and EP’s plane-fit LAT-gradient magnitude. Boxes show the median and interquartile range, whiskers extend to 1.5× the interquartile range, and points are individual studies (atrial n=8, ventricular n=9); the dashed line marks r=0 (no agreement). Brackets give the paired MCG-versus-ECGi contrast (two-sided Wilcoxon signed-rank): activation time p=0.38 (atrial) and 0.65 (ventricular); conduction velocity p=0.46 (atrial) and 0.10 (ventricular). Detailed results in **Tables 2-3**.

**Table 2:**
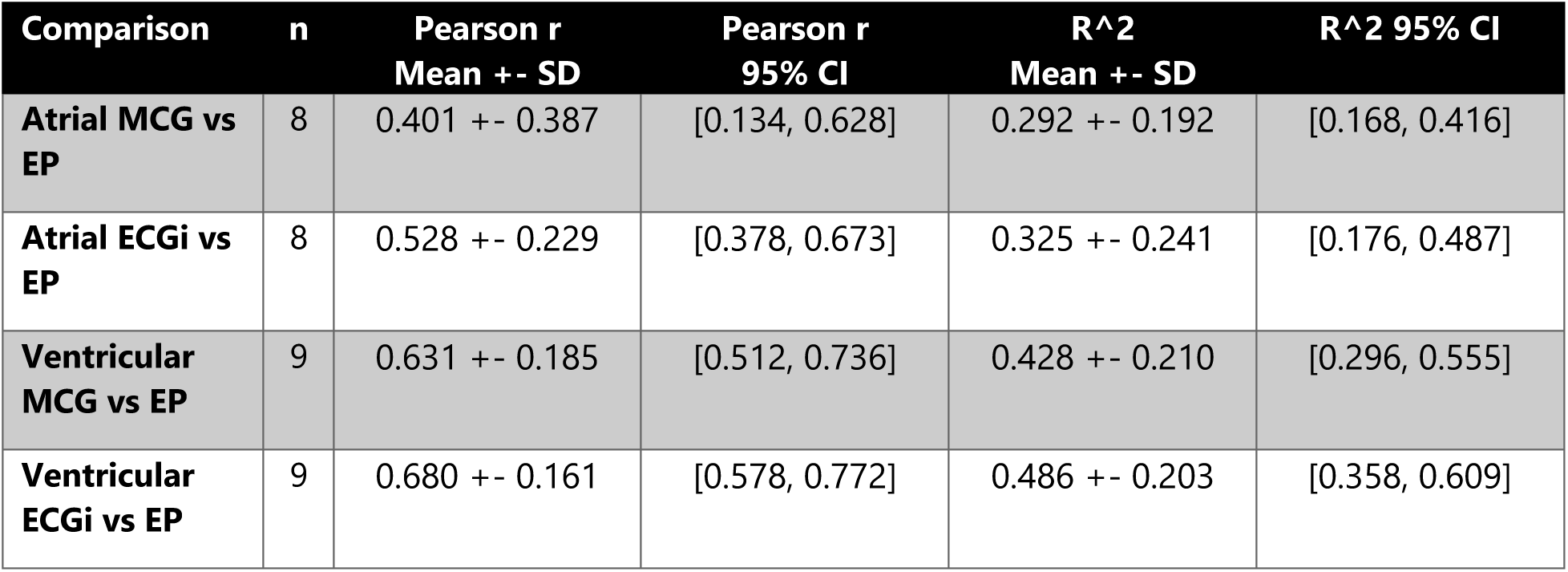
Activation-time agreement was computed on spatially smoothed, zero-referenced LAT maps; atrial analyses include 8 datasets and ventricular include 9. Spearman ρ and the paired MCG-vs-ECGi Wilcoxon signed-rank comparisons are reported in the text. Each study contributed a single agreement value per modality; bootstrap percentile intervals (10,000 resamples) and all paired comparisons were computed across these per-study values (n=8 atrial, n=9 ventricular). Resampling was performed at the subject level rather than over pooled vertices.

| Comparison | n | Pearson r<br>Mean +- SD | Pearson r<br>95% CI | R<sup>2</sup><br>Mean +- SD | R<sup>2</sup> 95% CI |
| --- | --- | --- | --- | --- | --- |
| Atrial MCG vs EP | 8 | 0.401 +- 0.387 | [0.134, 0.628] | 0.292 +- 0.192 | [0.168, 0.416] |
| Atrial ECGi vs EP | 8 | 0.528 +- 0.229 | [0.378, 0.673] | 0.325 +- 0.241 | [0.176, 0.487] |
| Ventricular MCG vs EP | 9 | 0.631 +- 0.185 | [0.512, 0.736] | 0.428 +- 0.210 | [0.296, 0.555] |
| Ventricular ECGi vs EP | 9 | 0.680 +- 0.161 | [0.578, 0.772] | 0.486 +- 0.203 | [0.358, 0.609] |

**Table 3:** Conduction-velocity agreement between each noninvasive modality and the invasive EP reference. Vertex-wise conduction velocity was estimated as the magnitude of a local least-squares plane-fit gradient of the spatially smoothed, zero-referenced LAT field and compared with the EP reference by Pearson r (R² is the squared Pearson r). Each study contributed a single agreement value per modality; bootstrap percentile intervals (10,000 resamples) and the paired MCG-vs-ECGi Wilcoxon signed-rank comparisons reported in the text were computed across these per-study values (n=8 atrial, n=9 ventricular), i.e., at the subject level rather than over pooled vertices.

| Comparison | n | Pearson r<br>Mean +- SD | Pearson r<br>95% CI | R<sup>2</sup><br>Mean +- SD | R<sup>2</sup> 95% CI |
| --- | --- | --- | --- | --- | --- |
| Atrial MCG vs EP | 8 | 0.122 +- 0.157 | [0.017, 0.220] | 0.037 +- 0.036 | [0.014, 0.060] |
| Atrial ECGi vs EP | 8 | 0.088 +- 0.167 | [-0.026, 0.188] | 0.032 +- 0.021 | [0.019, 0.046] |
| Ventricular MCG vs EP | 9 | 0.218 +- 0.083 | [0.169, 0.271] | 0.054 +- 0.038 | [0.032, 0.079] |
| Ventricular ECGi vs EP | 9 | 0.158 +- 0.115 | [0.097, 0.238] | 0.037 +- 0.056 | [0.011, 0.077] |

In ventricular datasets (n=9), agreement with EP was high for both modalities: MCG vs EP mean Pearson r 0.63 ± 0.19 (Spearman ρ 0.68) and ECGi vs EP mean Pearson r 0.68 ± 0.16 (ρ 0.69), with no significant difference between modalities (Wilcoxon signed-rank p=0.65) and both correlating with EP above chance (each p≤0.004). Overall, both modalities demonstrated higher LAT agreement in ventricular than atrial pacing, with comparable agreement between MCG and ECGi in each chamber.

Conduction-velocity agreement with EP was numerically higher for MCG than ECGi in the ventricle (Pearson r 0.22 ± 0.08 vs 0.16 ± 0.12; Wilcoxon signed-rank p=0.10) and modest and comparable in the atrium (r 0.12 ± 0.16 vs 0.09 ± 0.17; p=0.46; **Table 3**).

## Discussion

In a controlled preclinical large-mammal model, with invasive epicardial electrophysiology as a physiologic reference standard, this study demonstrates multimodality validation of a novel solid-state MCG system. Across chamber-level datasets, MCG produced quantifiable SOO estimates and measurable LAT agreement with epicardial EP reference maps, supporting technical feasibility for noncontact localization and activation characterization. There are many potential applications for this approach, including personalized phenotyping for precision medicine,^14^ guidance of stereotactic arrhythmia radioablation,^3^ and complementing recent descriptions of MRI-guided ablation.^15^

When compared with ECGi, chamber-specific performance patterns were observed. In atrial pacing, MCG localized the site of origin significantly closer to the reference lead than ECGi (paired Wilcoxon signed-rank p=0.023); in ventricular pacing, MCG showed a numerically lower median distance-to-lead error that did not reach statistical significance (p=0.074). For activation-time agreement on spatially smoothed maps, both modalities tracked the invasive EP reference well, with higher agreement in ventricular than atrial pacing, and there was comparable performance between MCG and ECGi within each chamber, where each correlated with EP above chance. Notably, MCG localized the atrial site of origin significantly closer to the reference lead than ECGi even though its atrial whole-field LAT agreement was only modest and more variable (reflecting two low-correlation atrial studies), suggesting that accurate origin localization does not require high global activation-time correlation. Conduction-velocity agreement was modest overall and numerically higher for MCG than ECGi in the ventricle, consistent with the greater spatial complexity of velocity than of activation timing.

These findings are consistent with the premise that magnetic-field-based reconstruction captures clinically relevant activation information without body-surface contact. The lower SOO error and higher activation pattern correlations in the ventricle are consistent with prior work in ECGi and are expected given the thinner-walled and more anatomically complex atrial regions.

Future developments of the work could implement artificial intelligence to improve SOO localization, which could include patient-specific cellular models of the atria and ventricles^16–18^ or identification of non-pulmonary vein triggers during atrial fibrillation.^13^

Several limitations should be noted. First, this was a controlled preclinical pacing model with modest sample size, and findings therefore require direct clinical validation. Second, the torso and heart geometry of the model increases the distance between sensors on the posterior aspect of the torso and creates more complex geometry near the sternum compared with humans. Third, endpoint derivation depends on preprocessing choices, including trim-mask definition and SOO cluster selection rules, which can influence absolute error estimates. These factors motivate larger prospective validation with prespecified analysis lock, and sensitivity analyses across reconstruction and registration settings.

These results support continued development of solid-state MCG as a practical noninvasive mapping modality. Immediate next steps include replication in larger and more heterogeneous cohorts and evaluation under spontaneous arrhythmia conditions. If validated, this approach may provide a scalable pathway for preprocedural localization and complementary activation mapping in patients for whom invasive mapping is limited or higher risk.

### Conclusions

In this preclinical large-mammal model, solid-state MCG was feasible for noninvasive site-of-origin localization, activation mapping, and exploratory conduction velocity reconstruction during both atrial and ventricular pacing. Median site-of-origin localization error was lower for MCG than for ECGi in both chambers, and MCG’s activation-time and conduction-velocity agreement with invasive EP mapping was comparable to that of ECGi. These findings establish technical feasibility and motivate larger prospective studies to define the generalizability and clinical role of solid-state MCG in noninvasive arrhythmia mapping.

## Abbreviations

CV: conduction velocity
ECGi: electrocardiographic imaging
EP: electrophysiology
LAT: local activation time
MCG: magnetocardiography
MRI: magnetic resonance imaging
SOO: site of origin

## Data Availability

The data that support the findings of this study are available from the corresponding author upon reasonable request.

## Author Contributions

Kelly A. Brennan and Sabyasachi Bandyopadhyay contributed equally to this work as co-first authors. Data collection, analysis, and algorithmic development were performed by Brennan KA, Bandyopadhyay S, and Sillett C. MCG system support and software development for MCG signal analysis and filtering were provided by Kameno M and Terazono Y. The ECGi system was developed by Rodrigo M, Brennan KA, Bandyopadhyay S, and Rogers AJ. Study protocol design and development were led by Rogers AJ, Narayan SM, Wang PJ, and Kameno M. Animal care and procedural support were provided by Lyons JK. MRI acquisition and sequence development were performed by Yang PC, Ikeda G, Takashima H, Matsura Y, and Ieki M. Critical manuscript revision and scientific feedback were provided by Ganesan P, Liu X, Wang PJ, Narayan SM, Rogers AJ, Brennan KA, and Bandyopadhyay S. All authors reviewed and approved the final manuscript.

## Acknowledgments

We acknowledge electrophysiology mapping support from Hana Jue, Hill Belfi, Logan Tarman, Tanner Filion, and Samana Shah.

## Supplementary Materials

### Supplementary Methods

#### (1) Mesh Harmonization and Vertex Correspondence

Cross-modality alignment was performed using rigid iterative closest point (ICP) registration with the EP mesh as reference geometry. After alignment, candidate one-to-one vertex correspondence was initialized via rounded-coordinate buckets and then completed with nearest-neighbor assignment for unresolved vertices, enforcing a bijective mapping. Mapping diagnostics (median distance, 95th-percentile distance, and maximum distance) were recorded to verify geometric correspondence quality before endpoint calculation.

#### (2) Flat-Region Trimming and Boundary Diagnostics

To avoid bias from open-boundary or nonphysiologic flat regions, a trim mask was generated on the EP reference mesh and propagated across modalities through the harmonization map. The cut-side plane was inferred using boundary-informed geometry when available, or low-curvature random sample consensus inference when explicit boundary edges were insufficient.

Normalized signed distance to the inferred plane was used to seed a contiguous flat-side component, from which an annulus-like boundary ring and region-growing rule defined the final removed region. The complementary keep mask defined the analyzable chamber surface.

#### (3) Earliest-Activation Cluster Construction

Earliest-activation candidates were selected from trimmed finite LAT values using the prespecified early-point rule. Mesh-adjacency connected-component analysis was then applied to isolate spatially coherent early-activation structure. The retained SOO component was summarized by centroid location, component cardinality, and optional spread descriptors.

---

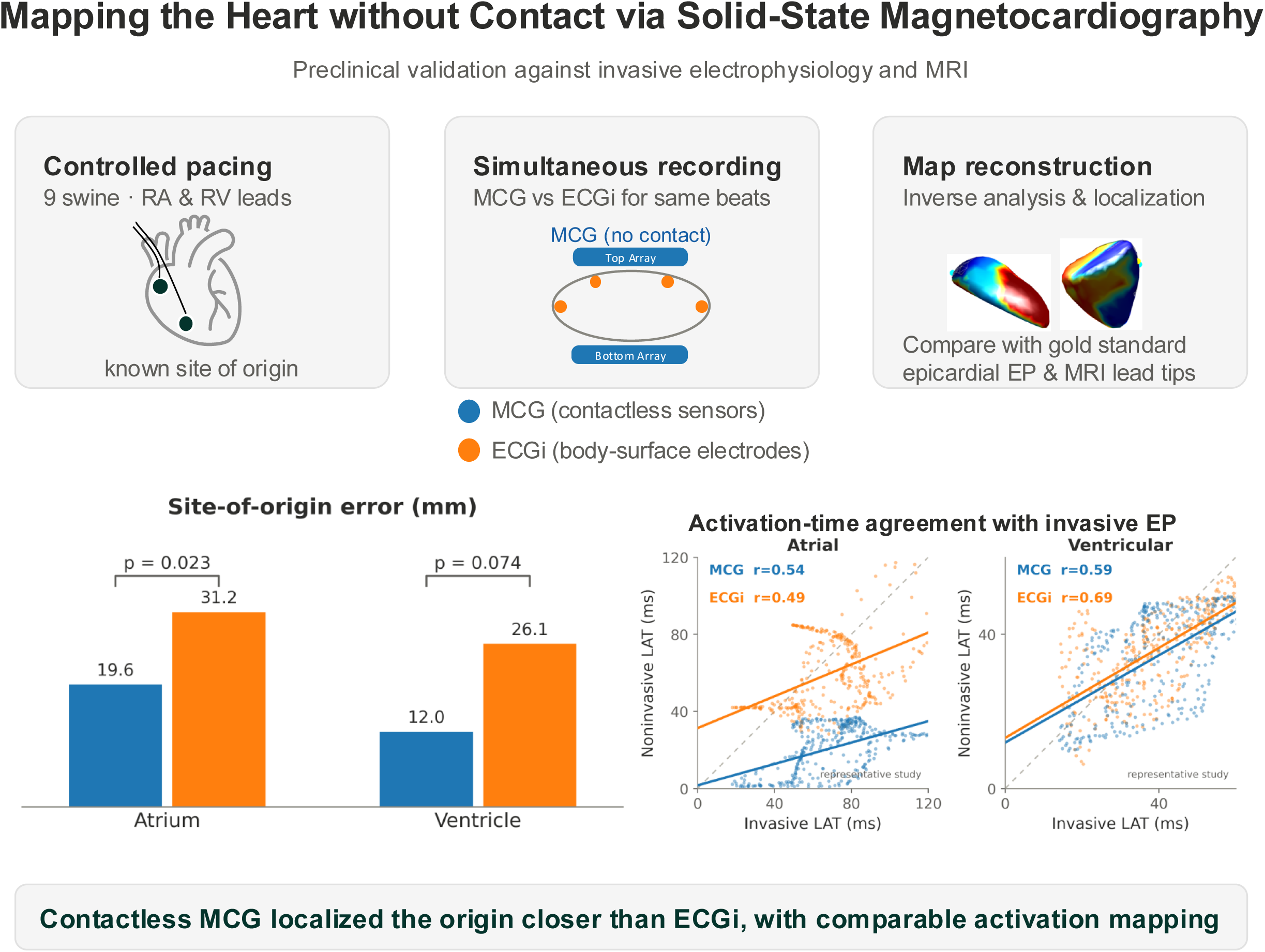

